# Detection of Human Bocavirus in Stool Samples Collected During an Outbreak of Acute Gastroenteritis in Children in Salvador, Bahia, Brazil

**DOI:** 10.64898/2026.10.21.26365923

**Authors:** Beatriz Peres Vidal, Silvana Oliveira Neves, Ively Paixão Santos, Silvia Ines Sardi, Gubio Soares Campos

## Abstract

Introduction: This study investigated human bocavirus (HBoV) in samples obtained during an outbreak of acute gastroenteritis (AGE) among children in Salvador, Bahia, Brazil. AGE is characterized by symptoms including diarrhea, vomiting, abdominal pain, and fever and may require hospitalization in severe cases. Methods: A total of 131 stool samples collected from children hospitalized with AGE at a hospital in Salvador, Bahia, were analyzed. HBoV was detected using nested polymerase chain reaction (nested PCR). Positive samples were sequenced and inoculated into Vero cell cultures, followed by sodium dodecyl sulfate– polyacrylamide gel electrophoresis (SDS-PAGE) and Western blotting. Results: Of the 131 samples analyzed, 13 tested positive for HBoV. Children aged 0–4 years were the most affected by AGE and showed the highest frequency of HBoV detection. HBoV was successfully cultured in Vero cells, representing a novel finding reported in this study. Subsequent SDS-PAGE and Western blot analyses revealed protein bands suggestive of HBoV structural proteins. Conclusion: This study highlights the need for accurate diagnostic methods and preventive strategies to control the spread of HBoV and other pathogens associated with gastroenteritis in pediatric populations.

## Introduction

Acute gastroenteritis is common among children and may lead to hospitalization in severe cases. It is characterized by inflammation of the gastrointestinal tract, with symptoms including diarrhea, vomiting, abdominal pain, and fever (Cheng et al., 2008). Human bocavirus has been investigated among the possible viral agents associated with this condition (Lin et al., 2020).

Human bocavirus (HBoV) was discovered in 2005 by Allander and colleagues in Sweden (Allander, 2008), following genomic investigations of respiratory samples from children with respiratory illness (Bastien et al., 2007; Mohammadi, 2023). HBoV is a single-stranded DNA virus belonging to the family Parvoviridae, which has been divided into the subfamilies Parvovirinae and Densovirinae (Nora-Krukle et al., 2018). The subfamily Parvovirinae includes viruses infecting animals and has been described as comprising eight genera: Amdoparvovirus, Aveparvovirus, Copiparvovirus, Dependoparvovirus, Erythroparvovirus, Protoparvovirus, Tetraparvovirus, and Bocaparvovirus. HBoV belongs to the genus Bocaparvovirus, which includes viruses infecting several mammals, including humans (Lin et al., 2020).

Four HBoV genotypes have been described. HBoV-1 is predominantly associated with respiratory disease, whereas HBoV-2, HBoV-3, and HBoV-4 have been associated with gastrointestinal disease. Under the classification cited by the authors, HBoV-1 and HBoV-3 belong to the species Primate bocaparvovirus 1, while HBoV-2 and HBoV-4 belong to Primate bocaparvovirus 2 (ICTV, 2017).

The HBoV genome consists of linear single-stranded DNA (ssDNA), which may have either positive or negative polarity and contains approximately 5,300 nucleotides. The virus encodes nonstructural proteins NP1, NS1, NS1-70, NS2, NS3, and NS4 (Shen et al., 2015), as well as structural proteins VP1 and VP2 (Li et al., 2015). The virion is nonenveloped and approximately 18–26 nm in diameter, with an icosahedral capsid described as containing 60 copies of VP1 and VP2 proteins (Guido et al., 2016).

The replication cycle of human bocavirus has not yet been fully elucidated. Nevertheless, several aspects of its replication have been described, including mechanisms shared with other members of the family Parvoviridae. The 3′ and 5′ termini of the HBoV genome contain palindromic sequences that form hairpin structures essential for viral DNA replication (Huang et al., 2012). Within the nucleus, viral DNA serves as a template for the host-cell replication machinery, including cellular DNA polymerase. A double-stranded intermediate is formed and cleaved by a viral endonuclease, probably NS1, generating free 3′-OH ends for continued replication (Berns & Parrish, 2013; Astell et al., 1985). Capsid proteins VP1 and VP2 assemble in an ordered fashion and encapsidate newly replicated viral genomes to generate infectious viral particles (Berns & Parrish, 2013). Viral assembly occurs in the host-cell nucleus, where structural components and viral genomes are assembled into mature particles (Santos, 2021). The newly assembled capsids containing viral genomes are subsequently released from host cells, possibly through cell lysis, completing the HBoV replication cycle (Berns & Parrish, 2013).

HBoV has been reported in association with acute gastroenteritis and respiratory infections. Given its respiratory tropism, HBoV infection has been associated with severe respiratory illnesses, including pneumonia and bronchiolitis (Mohammadi, 2023). Other reported clinical conditions include acute conjunctivitis (Johnson et al., 2023), myocarditis (Thanasia et al., 2023), and chronic otitis media (Runge et al., 2023).

Epidemiological studies conducted in several regions of the world have detected HBoV in stool samples from patients with acute gastroenteritis. Reported detection frequencies vary substantially, ranging from 2.3% to 50.6% across the studies cited (Bergallo et al., 2018; Cheng et al., 2008; Mohammad et al., 2020; Nora-Krukle et al., 2018). In Brazil, a study including 705 children reported HBoV detection in 2% of participants (Carolina et al., 2007). Another investigation in Arraial do Cabo, Rio de Janeiro, was cited as reporting HBoV in 53.8% of the samples evaluated (do Nascimento et al., 2022). These studies are discussed as evidence of the geographical distribution of HBoV and its possible relevance to gastrointestinal infection in children.

Beyond differences in tissue tropism, genetic variation has been proposed as a factor contributing to the association of HBoV with both gastrointestinal and respiratory disease. Although HBoV is a DNA virus, its genome can undergo mutations. It has therefore been hypothesized that an enteric HBoV lineage may have given rise to respiratory HBoV-1 (Lekana-Douki et al., 2018).

HBoV continued to be detected in patients during the COVID-19 pandemic, despite changes in the circulation of respiratory viruses during that period (Lai et al., 2022). A study comparing hospitalizations for bronchiolitis before and after the onset of COVID-19 also described an increase in cases associated with bocavirus in France (Berdah et al., 2022).

## Materials and Methods

This descriptive cross-sectional study was approved by the Research Ethics Committee of the Institute of Health Sciences, Federal University of Bahia (ICS-UFBA), under CAAE registration number 61239422.9.0000.5662. A total of 131 stool samples from an acute gastroenteritis outbreak in Salvador, Bahia, Brazil, between May and September 2024 were analyzed. Following collection, samples were transported under refrigeration to the ICS-UFBA Virology Laboratory and stored at −80 °C until HBoV testing.

For PCR testing, the samples were processed for nucleic acid extraction. Briefly, 300 µL of stool were suspended in 1 mL of nuclease-free water, homogenized for 15 seconds, and centrifuged at 8,000 rpm for 1 minute. Nucleic acids were subsequently extracted from 140 µL of the resulting supernatant using the QIAamp Viral DNA Kit® (QIAGEN) according to the manufacturer’s instructions.

Human bocavirus was investigated using nested PCR according to the protocol reported by Kapoor et al. (2011). GoTaq® G2 Colorless Master Mix (Promega) was used for amplification. An HBoV-2 strain previously detected in Salvador and deposited in GenBank under accession number KM366092 was included as a positive control. Amplicons were visualized by electrophoresis on 2% agarose gels stained with ethidium bromide and examined under ultraviolet illumination. The expected HBoV amplicon was 576 base pairs long.

Virus isolation experiments were performed using Vero CCL-81 cells obtained from the American Type Culture Collection (ATCC). Cells were maintained in Dulbecco’s Modified Eagle Medium (DMEM) supplemented with 10% fetal bovine serum, penicillin, streptomycin, and amphotericin B. Cultures were incubated at 37 °C in a humidified atmosphere containing 5% CO_2_.

Suspensions for cell inoculation were prepared from HBoV-positive stool samples. One gram of stool was diluted in 5 mL of phosphate-buffered saline (PBS) supplemented with penicillin, streptomycin, and amphotericin B. After vortex mixing for 15 seconds, the material was centrifuged at 7,700 rpm for 20 minutes at 4 °C. The resulting supernatant was used as the viral inoculum.

Two procedures were used to inoculate Vero cells. In the first, based on Yamashita et al. (1991), 500 µL of inoculum were combined with 500 µL of freshly trypsinized cells at a concentration of 5 × 10^5^ cells/mL. The mixture was incubated at 37 °C for 1 hour with agitation every 10 minutes to facilitate viral adsorption. It was then centrifuged at 960 rpm for 5 minutes at 4 °C. The supernatant was removed, and the cell material was transferred into 12-well plates containing 3 mL of DMEM supplemented with 2% fetal bovine serum, antibiotics, and an antifungal agent.

For the second procedure, based on Cann (1999), Vero cell monolayers were grown for 24 hours in 12-well plates before inoculation. The culture medium was removed, and 500 µL of each sample were added directly to the monolayer. After 1 hour of viral adsorption, DMEM supplemented with penicillin and streptomycin was added. In both procedures, cultures were examined daily for up to seven days under an inverted microscope to monitor morphological changes and possible cytopathic effects. Infection was assessed by molecular testing.

Immunofluorescence was performed on Vero cultures inoculated with HBoV. Cells were dispensed onto multiwell microscope slides (25 µL per well), dried in an incubator, and fixed with acetone for 10 minutes. The slides were then incubated at 4 °C with serum obtained from a patient after HBoV infection, diluted 1:10. After three washes with PBS, fluorescein isothiocyanate (FITC)-conjugated anti-human IgG secondary antibody specific to the Fab region was added at a dilution of 1:1,000. Slides were incubated for 1 hour, washed three more times with PBS, and counterstained with Evans blue for 3 minutes before examination by fluorescence microscopy.

For characterization of antigenic proteins, a viral suspension of strain 308 was obtained from Vero cell cultures maintained in 300-mL Roux flasks. After freeze–thaw cycles, the material was centrifuged at 10,000 rpm at 10 °C. The supernatant underwent ultracentrifugation at 100,000 × g, and the resulting pellet was resuspended in 300 µL of concentrated NET buffer. The preparation was stored at −80 °C until protein characterization assays were performed.

Proteins were separated by sodium dodecyl sulfate–polyacrylamide gel electrophoresis (SDS-PAGE), following Laemmli (1970). Concentrated viral preparations and uninoculated Vero-cell controls were analyzed. Before loading, samples were heated to 100 °C for 3 minutes in denaturing buffer. Electrophoresis was carried out in a 12% polyacrylamide gel at 100 V. The gel was then fixed for 1 hour in a solution containing 10% acetic acid and 40% methanol, stained with 0.25% Coomassie Brilliant Blue R-250 for 24 hours, and destained under agitation using 5% acetic acid and 10% methanol until protein bands became visible.

Molecular weights were estimated using a Prestained Protein Ladder spanning 10–180 kDa (Abcam®, Cambridge, MA, USA). Migration distances were measured to calculate relative mobility (Rf), and a calibration curve based on the molecular-weight standard was used to estimate the sizes of the protein bands in the samples.

Protein detection was performed by Western blotting. Following SDS-PAGE, proteins were transferred to nitrocellulose membranes by electrotransfer at 100 V for 1 hour at 4 °C. Membranes were rinsed with distilled water and blocked with PBS containing 5% milk for 90 minutes at 25 °C. They were subsequently incubated for 90 minutes at 37 °C with serum from a patient with confirmed prior HBoV infection, diluted 1:10 in PBS containing 0.5% milk. After three 10-minute washes with 0.05% PBS-Tween under agitation, the membranes were incubated with horseradish peroxidase (HRP)-conjugated goat anti-human IgG (H+L), diluted 1:1,000 in the same diluent. Following a further 90-minute incubation at 37 °C, membranes were washed three times with 0.05% PBS-Tween and once with PBS (1×). Immunoreactive bands were visualized using diaminobenzidine (DAB) dissolved in PBS with hydrogen peroxide (H_2_O_2_).

## Results

### 1. Molecular detection of HBoV

Among 131 study samples analyzed by nested PCR, 13 (9.9%) tested positive for HBoV and 118 (90.1%) tested negative. PCR products were evaluated by agarose gel electrophoresis, which revealed the expected 576-bp amplicon (Figure 1).

**Figure 1.**
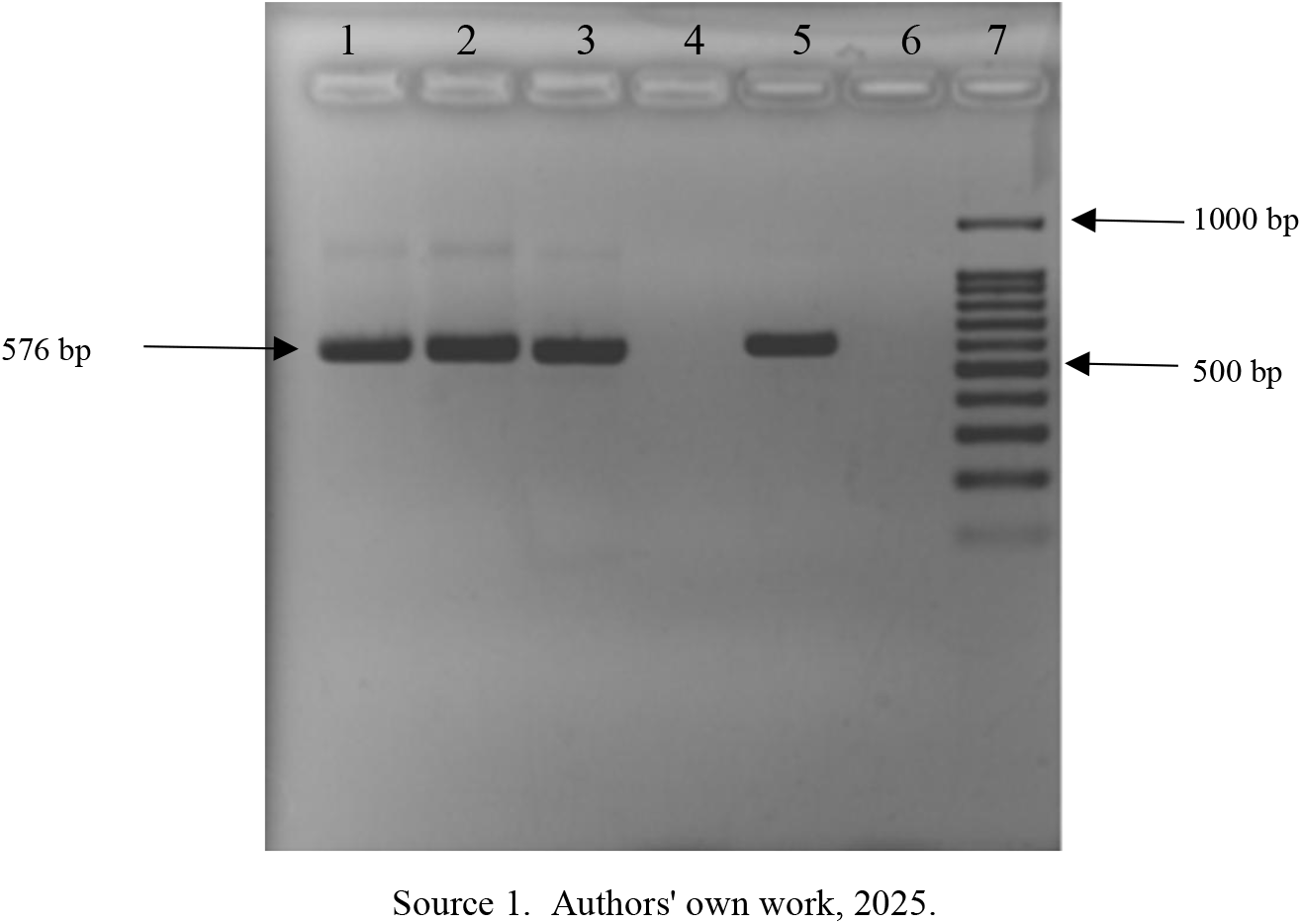
Detection of HBoV by nested PCR. Ethidium bromide-stained 2% agarose gel showing HBoV nested-PCR products. Lanes 1–3, HBoV-positive samples; lane 5, HBoV positive control; lanes 4 and 6, HBoV-negative samples; lane 7, 100-bp DNA ladder (Invitrogen, USA).

### 2. Age distribution of HBoV-positive cases

Most cases were observed among children aged 0–4 years, although the study included children up to 14 years of age (Table 1).

**Table 1.** Age distribution of HBoV-positive patients. The table shows HBoV-positive cases (n = 13) across age categories.

| Age (Years) | HBoV positive patients |
| --- | --- |
| 0-4 | 12 |
| 5-9 | 0 |
| 10-14 | 1 |

Source 2. Authors' own work, 2025.

### 3. Phylogenetic analysis

Sequence alignment and phylogenetic analyses were conducted using the maximum likelihood method in MEGA11. Three HBoV-1 strains and one HBoV-2 strain were identified. The HBoV-2 sequence showed the greatest similarity to Chinese strains FJ375127.1 and FJ375128.1 from 2008 (Figure 2), whereas HBoV-1 sequences clustered near strains from China and Argentina (Figure 3).

**Figure 2.**
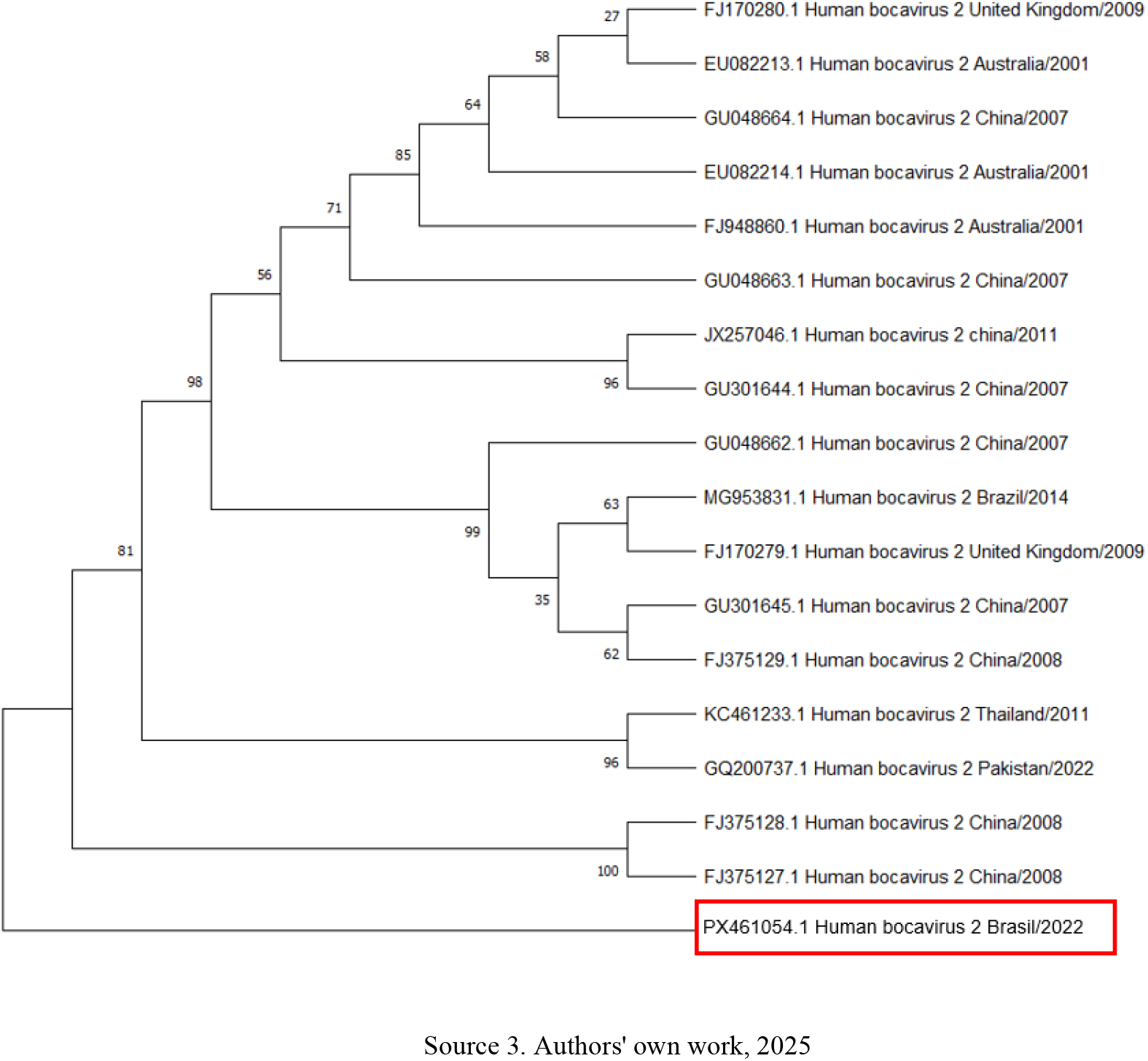
Phylogenetic analysis of the VP1/VP2 coding region. The tree was constructed using the composite maximum likelihood method, with branch lengths expressed as the number of nucleotide substitutions per site and 1,000 bootstrap replicates. The analysis included 18 HBoV-2 nucleotide sequences retrieved from GenBank (http://www.ncbi.nlm.nih.gov). Evolutionary analyses were conducted using MEGA11.

**Figure 3.**
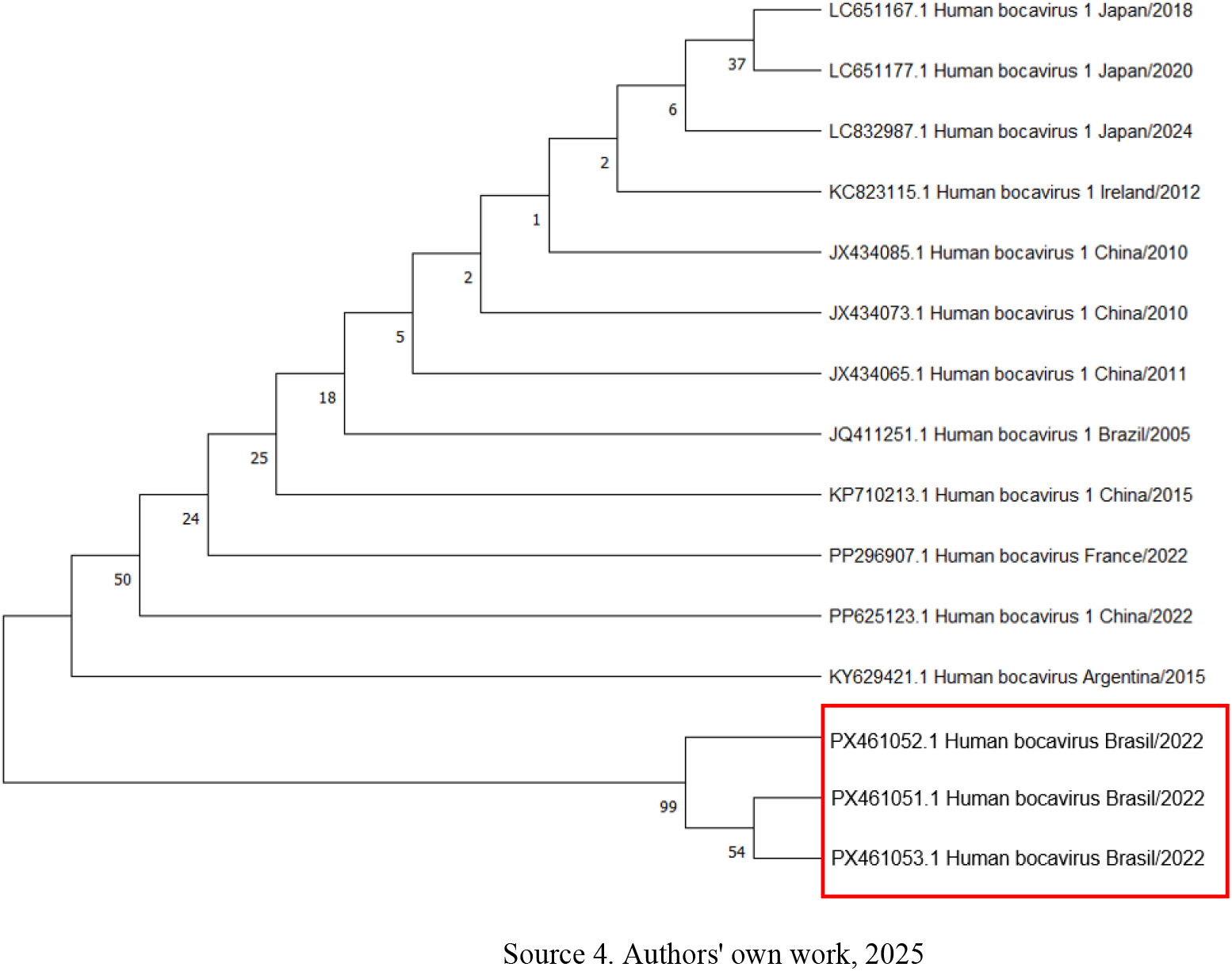
Phylogenetic analysis of the VP1/VP2 coding region. The tree was constructed using the composite maximum likelihood method, with branch lengths expressed as the number of nucleotide substitutions per site and 1,000 bootstrap replicates. The analysis included 15 HBoV-1 nucleotide sequences retrieved from GenBank (http://www.ncbi.nlm.nih.gov). Evolutionary analyses were conducted using MEGA11.

### 4. Viral isolation

All samples positive for HBoV by PCR were inoculated into Vero cell cultures. The cytopathic effects observed included cell rounding, fusion, and cell death (Figure 4), and viral isolation was reported as successful for two samples. Both inoculation approaches co-incubation with freshly trypsinized cells and inoculation of preformed monolayers were described as successful (see Materials and Methods). A viral titer of 1 × 10^6.5 TCID50/mL was reported for HBoV after 48 hours of observation.

**Figure 5.**
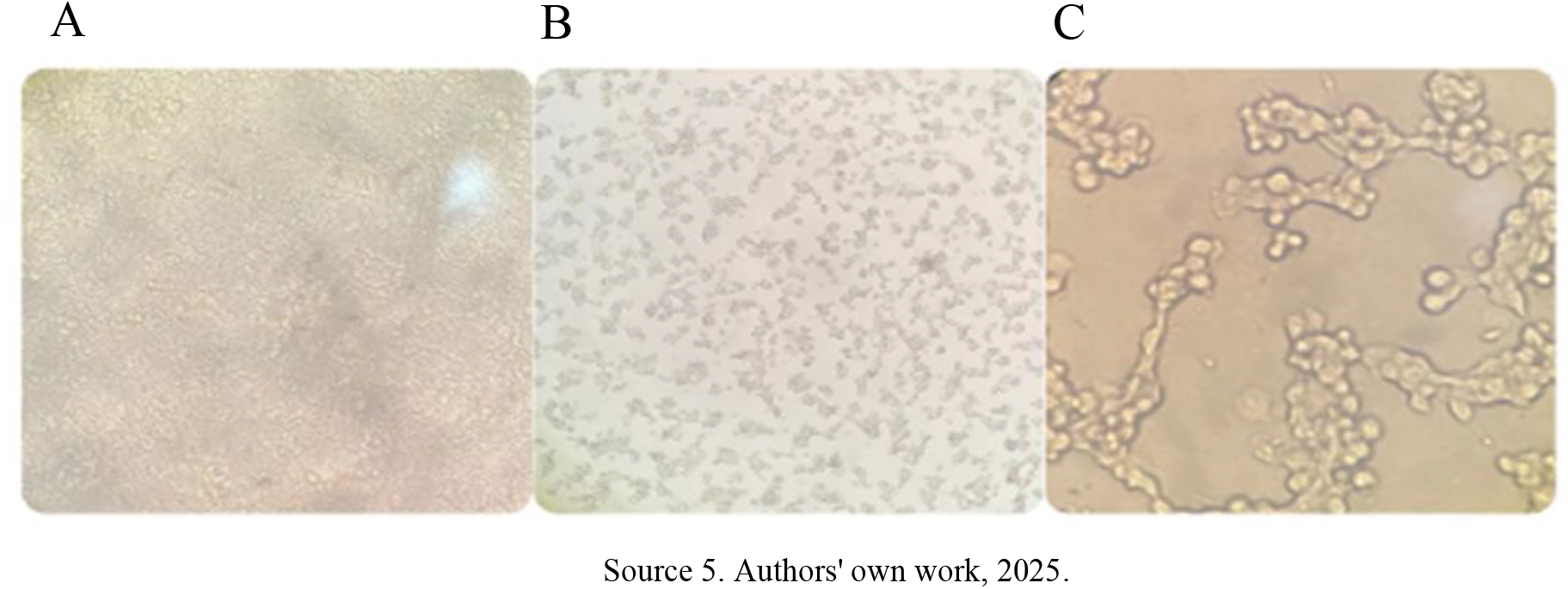
Light microscopy of cytopathic effects in Vero cells 48 hours after HBoV inoculation. (A) Uninfected Vero cells (negative control) showing normal cell morphology. (B) HBoV-inoculated Vero cells at 10× magnification. (C) HBoV-inoculated cells at 40× magnification.

### 7. Characterization of HBoV antigenic proteins

#### 7.1 SDS-PAGE and Western blot analysis

Western blotting (Figure 5) revealed three immunoreactive bands recognized by serum from a previously infected patient. The observed proteins were described as potentially corresponding to the HBoV structural proteins VP1, VP2, and VP3. Reported molecular masses are approximately 80–86 kDa for VP1, 64–75 kDa for VP2, and 60–62 kDa for VP3, the latter described as the most abundant capsid protein (Knipe & Howley, 2013). VP2 is also discussed as an important target of the host immune response.

**Figure 5.**
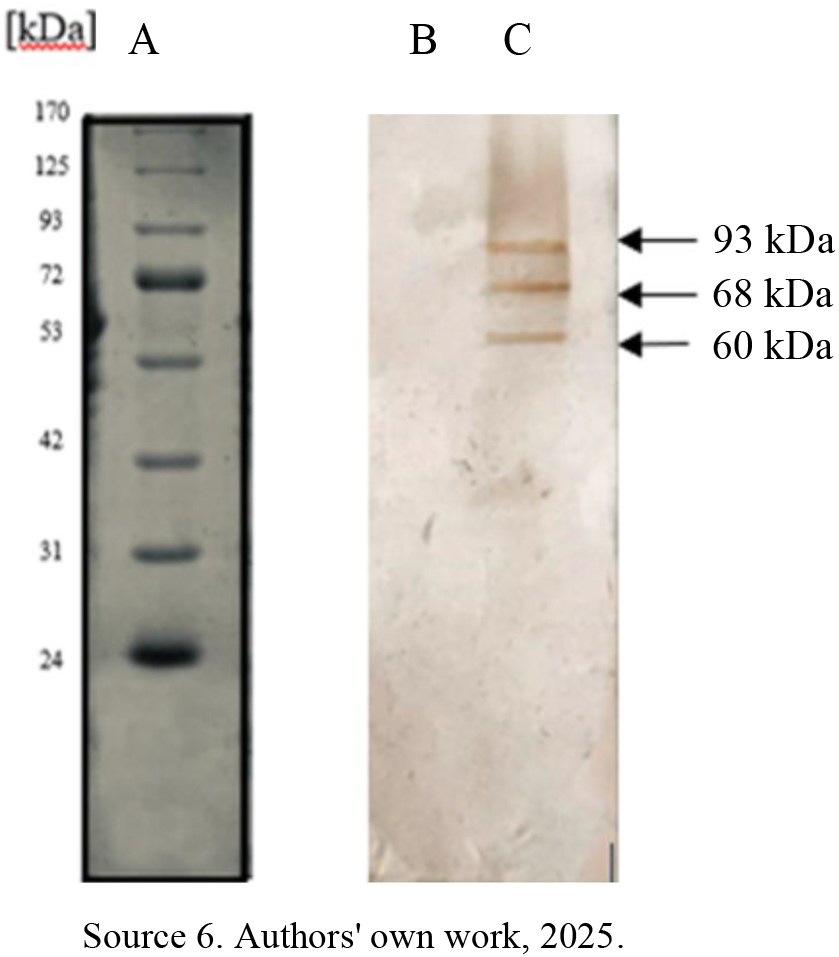
Western blot analysis following 12% polyacrylamide gel electrophoresis. (A) Prestained Protein Ladder, mid-range molecular weight (10–180 kDa; Abcam®, Cambridge, MA, USA). (B) Control sample with serum from a patient previously exposed to HBoV. (C) HBoV-inoculated cell preparation probed with patient serum. Bands are annotated in the original image at approximately 93, 68, and 60 kDa.

## Discussion

Acute gastroenteritis affects all age groups, although children and older adults are particularly affected. Common viral pathogens include norovirus, rotavirus, and human adenovirus; less extensively studied viruses, such as human bocavirus, have also been detected in association with gastroenteritis.

The findings indicate that HBoV continues to be detected among children with AGE in Salvador, as in an earlier study (Campos et al., 2016). HBoV has been reported in several countries, including in cases involving its detection alone and in coinfections with other pathogens (Cheng et al., 2008; Lekana-Douki et al., 2018; Lin et al., 2020; Nora-Krukle et al., 2018). Its widespread geographical distribution and genetic diversity underscore the need for surveillance and further studies to better characterize transmission, genetic variation, and potential public-health implications.

The detection of HBoV sequences genetically similar to strains reported in other countries was interpreted as evidence of viral circulation across national borders (Gaudin & Goetz, 2021). Distinct Brazilian phylogenetic clusters may reflect local selective pressures, mutations, or introductions of new lineages that may influence viral evolution and spread (Gogarten & Townsend, 2005; Sanjuán & Domingo-Calap, 2016).

Phylogenetic analysis can contribute to understanding viral evolution and may inform investigations into transmission, recombination, and mutations associated with pathogenicity. Further studies of circulating lineages and the environmental factors influencing HBoV evolution could support the development of diagnostic approaches and, potentially, preventive measures including virus-specific vaccines.

Among HBoV-positive samples, two were successfully isolated, with cytopathic effects observed after seven days and the presence of HBoV confirmed by PCR and sequencing. Earlier studies have described difficulties in culturing HBoV (Allander, 2008; Huang et al., 2012; Schildgen et al., 2008). HBoV has also been cultured in human colorectal adenocarcinoma (Caco-2) cells using specimens from patients with gastroenteritis (Ghietto et al., 2017). The authors did not identify previous published reports of HBoV isolation in Vero cells and therefore describe their finding as the first such report. They also consider the cytopathic effects observed to be previously undescribed.

Immunoreactive bands compatible with the putative HBoV capsid proteins VP1, VP2, and VP3 were observed in inoculated cells (Knipe & Howley, 2013). Western blotting revealed a stronger band compatible with VP2, which was interpreted as indicating a more pronounced immune response to this protein than to the others. This finding was compared with previous reports suggesting that VP2 is an important antigenic target (Deng et al., 2014).

Capsid proteins VP1 and VP2 have roles in HBoV–host interactions. The authors describe VP1 as participating in receptor binding and cell entry (Kantola et al., 2008; Ros et al., 2020). VP2 is discussed as a major HBoV antigenic determinant that stimulates an IgG antibody response relevant to virus neutralization and protection against reinfection (Bhat & Almajhdi, 2021).

The most intense band in the Western blot was compatible with VP2. The authors interpreted this observation as supporting a stronger response to VP2, as described in the literature, but acknowledge that additional analyses are required to confirm the identity of the protein (Bhat & Almajhdi, 2021). This observation was also discussed in relation to VP2 immunogenicity and its potential relevance to vaccine development (Deng et al., 2014). In addition, presentation of VP2-derived antigens to CD4+ T lymphocytes may induce cytokine production, including IFN-γ, influencing innate and adaptive immunity and immunological memory (Bhat & Almajhdi, 2021). Further research is required to clarify these immune mechanisms and investigate VP2 as a potential vaccine target.

These findings may inform future research on viral antigens and the development of more specific diagnostic assays for HBoV. Characterization of capsid proteins may also contribute to the design of vaccine-related studies aimed at reducing the burden of infection in vulnerable populations. The authors therefore regard these observations as potentially relevant to public health.

Following an analysis of the age distribution of viral acute gastroenteritis (AGE) cases among children, those aged 0–4 years were found to be the most affected in the study population. These findings suggest that younger children are more frequently affected by AGE, including cases of viral acute gastroenteritis caused by human bocavirus (HBoV). Furthermore, studies conducted in Brazil have shown that children under three years of age accounted for the highest number of deaths due to diarrhea and gastroenteritis of infectious origin in the country between 2017 and 2022 (De Melo et al., 2023).

Several factors may contribute to the burden of gastroenteritis in young children, including immunological immaturity (Shaw & Su, 2021), exposure to previously unencountered pathogens, and greater susceptibility to dehydration. Severe diarrhea may cause dehydration (Brandt et al., 2015), placing young children at increased risk of complications (Pijnacker et al., 2019). HBoV has also been described in association with respiratory infection (Bagasi et al., 2020), myocarditis (Thanasia et al., 2023), and severe disease requiring intensive care (Liao et al., 2022). Monitoring the circulation of viral agents in pediatric populations may therefore have clinical and epidemiological relevance.

Viral isolation from stool presents major challenges because fecal specimens contain diverse microorganisms and a complex biological matrix (Cao et al., 2022; Eckburg et al., 2005). Bacteria, fungi, and other agents may interfere with the isolation and propagation of the target virus and increase the risk of contamination during laboratory procedures.

The viral isolation results provide a basis for further investigation of the biological properties of these viruses. The discussion also refers to possible future studies of human adenovirus (HAdV) and HBoV in cell culture (Wang, 2020). Both isolation strategies are described as suitable for the objectives of this study, although further experiments would be needed to compare their performance.

Viral cultivation and isolation are important tools in virology, enabling molecular and functional analyses of viruses and their interactions with host cells. These approaches may support diagnostic assay development, vaccine research, and the characterization of viral proteins (Mello, 2010; Silva et al., 2021), including SDS-PAGE and Western blotting. Cell-culture experiments may also be combined with functional assays and fluorescence-based techniques to investigate infection, viral replication, and virus–host protein interactions (Zhang & Gauger, 2014; Aoki et al., 2010).

## Conclusions

This study confirmed the detection of human bocavirus (HBoV) in pediatric acute gastroenteritis cases in Bahia, Brazil. Molecular and phylogenetic characterization revealed genetic diversity among the HBoV strains detected in Brazil, including sequences closely related to lineages reported in other countries. These observations support further investigation of HBoV evolution and dissemination. The higher number of AGE cases among children aged three years or younger also emphasizes the importance of monitoring viral agents in this population, given their greater vulnerability to clinical complications.

HBoV isolation in Vero cells was reported as confirmed by nested PCR and sequencing of part of the viral genome. Successful isolation is potentially valuable for further investigation of viral characteristics. Western blotting revealed immunoreactive bands assigned to putative viral proteins, with the most intense band considered compatible with VP2, offering directions for future research into HBoV antigenicity, diagnostics, and vaccine-related strategies.

Overall, the study highlights the use of viral culture, molecular detection, phylogenetic methods, and immunological techniques to advance research into HBoV. The findings may provide a foundation for further virological studies and improved epidemiological surveillance.

## Data Availability

All data produced in the present work are contained in the manuscript

